# A noninvasive clinical method to measure *in vivo* mechanical strains of the lamina cribrosa by optical coherence tomography

**DOI:** 10.1101/2023.08.14.23294082

**Authors:** Vanessa Hannay, Cameron A. Czerpak, Harry A. Quigley, Thao D. Nguyen

**Author notes:** **Corresponding Author:** Harry A. Quigley MD, 900 Bellemore Road, Baltimore, Maryland USA 21210.

## Abstract

**Objective:** To measure mechanical strain of the lamina cribrosa (LC) after intraocular pressure (IOP) change produced one week after a change in glaucoma medication.

**Design:** Cohort study.

**Participants:** Adult glaucoma patients (23 eyes, 15 patients) prescribed a change in IOP-lowering medication.

**Intervention:** Non-invasive optical coherence tomography (OCT) imaging of the eye.

**Main Outcomes:** Deformation calculated by digital volume correlation of OCT scans of the LC before and after IOP lowering by medication.

**Results:** Among 23 eyes, 17 eyes of 12 persons had IOP lowering ≥ 3 mmHg (reduced IOP group) with tensile anterior-posterior *E*_*zz*_ strain = 1.0% ± 1.1% (p = 0.003) and compressive radial strain (*E*_*rr*_) = -0.3% ± 0.5% (p=0.012; random effects models accounting inclusion of both eyes in some persons). Maximum in-plane principal (tensile) strain and maximum shear strain in the reduced IOP group were: *E*_*max*_ = 1.7% ± 1.0% and *Γ*_*max*_ = 1.4% ± 0.7%, respectively (both p<0.0001 versus zero). Reduced IOP group strains *E*_*max*_ and *Γ*_*max*_ were significantly larger with greater %IOP decrease (<0.0001, <0.0001). The compliance of the *E*_*zz*_, *E*_*max*_, and *Γ*_*max*_ strain response, defined as strain normalized by the IOP decrease, were larger with more abnormal perimetric mean deviation or visual field index values (all p≤0.02). Strains were unrelated to age (all p≥0.088). In reduced IOP eyes, mean LC anterior border posterior movement was only 2.05 μm posteriorly (p=0.052) and not related to % IOP change (p=0.94, random effects models). Only *E*_*rr*_ was significantly related to ALD change, becoming more negative with greater posterior LC border change (p=0.015).

**Conclusion:** LC mechanical strains can be effectively measured by changes in eye drop medication using OCT and are related to degree of visual function loss in glaucoma.

**Trial Registration:** ClinicalTrials.gov Identifier: NCT03267849

## Introduction

Glaucoma is the second most common cause of blindness worldwide^1^ and numerous studies show that lowering intraocular pressure (IOP) slows its progressive vision loss.^2^ The level of IOP is a major risk factor for glaucomatous damage, whether IOP is within or above the normal range.^3^ Methods to lower IOP include surgery, laser treatment, and eye drop medication. While modest IOP lowering leads to stable visual function in many glaucoma patients, a minority have a much more aggressive course.^4^ At present there are not consistent biomarkers that indicate which eyes will have more rapid progression with standard initial therapy.

Glaucomatous neuropathy results from death of retinal ganglion cells, due in part to injury to their axons at the optic nerve head (ONH) within its lamina cribrosa (LC).^5^ The upper and lower zones of the LC, through which the most susceptible axons pass, have been shown to have less dense connective tissue structure and to undergo greater mechanical strain with *in vitro* inflation testing.^6^ This suggests that a clinical measure of LC strain might provide valuable information on which patients and what LC zones would have greater susceptibility to rapid progressive worsening.^7,8^ We and others have measured the short-term strain responses of the LC in glaucoma patients after both increases and decreases in IOP, using digital volume correlation of spectral domain (SD)--optical coherence tomography (OCT) images taken at two different IOPs.^9,10,11,12,13,14^ With modest IOP lowering, biomechanical strains are regularly obtainable and fit with the expected deformation of the LC tissue, undergoing expansion in the anterior—posterior direction and contraction in diameter.^15^ Artificial elevation of IOP produced corresponding compression of the lamina. Most importantly, our data and that of Girard and co-workers indicate that eyes with greater loss of RGC have greater LC strain.^15,16^ These data suggest that short-term LC strain measurements may provide biomarkers for glaucoma susceptibility.

In the present report, we applied digital volume correlation (DVC) to analyze OCT images of the LC in glaucoma patients before and one week after a change of IOP-lowering eye medication to calculate the strains in the LC. Our purpose was to produce a method for clinical strain assessment that would be compatible with standard management approaches in initial glaucoma therapy for any office treating patients that have OCT capability.

## Methods

### Experimental subjects

The study was approved by the Johns Hopkins Institutional Review Board and written informed consent was obtained from all subjects, who were patients from Glaucoma Center of Excellence, Wilmer Ophthalmological Institute, Johns Hopkins Medicine. Data are presented for 23 eyes of 15 patients. Data from additional patients were excluded due to poor image quality. Among the 23 eyes, all but two were from patients starting hypotensive eye drops, while one eye was from a patient instructed to stop eye drops, while the final patient started oral glaucoma medication.

Eyes were imaged by OCT (Spectralis, Heidelberg Engineering, Heidelberg, Germany) before and 7.3 ± 1.4 days after IOP change, using methods described previously.^15^ Prior to the first imaging session, all eyes underwent a standard glaucoma examination to assess disease progression, which measured their average retinal nerve fiber layer (RNFL) thickness and visual field status (Zeiss Humphrey Field Analyzer (HFA2i or 3) 24-2 Swedish Interactive Threshold Algorithm (SITA Standard) field tests; Zeiss HFA2i, Dublin, CA) with indices mean deviation (MD) and visual field index (VFI) within 3 months of imaging. Retinal nerve fiber layer (RNFL) thickness was measured using Cirrus OCT (Zeiss). The average MD for the overall group of 23 eyes was -5.4 dB ± 5.7 dB, with 15 eyes having MD > -6 dB, 5 eyes with moderate damage (−12 dB < MD < -6 dB), and 3 eyes with severe damage (MD < -12 dB). The average thickness of the retinal nerve fiber layer (RNFL) ranged from 54 to 83 μm. The age range of the 15 study subjects was 30 to 80 years (mean = 62.1 ± 12.4 years; median, 64 years).

We divided the 23 eyes into reduced IOP and unchanged IOP groups, based on whether or not they underwent significant IOP change. Reduced IOP eyes (n=17 eyes, 12 patients) was defined as eyes with an IOP decrease ≥3 mmHg, while unchanged IOP eyes (6 eyes, 5 patients) had ≤1 mmHg change. None of the eyes had IOP change between 1 and 3 mmHg. The one patient who stopped eye drops to attempt a change in IOP had no change and is assigned to the unchanged IOP group. The reduced IOP group had a higher average baseline IOP (20.3±5.8 mmHg) than the unchanged IOP group (12.5±5.2 mmHg), but the two groups did not differ in the average age, MD, VFI, or RNFL thickness.

### Optical coherence tomography imaging

IOP was measured using a rebound tonometer (iCare Finland Oy) directly before each imaging session. Baseline and post-treatment IOP were each recorded as the mean of six consecutive measurements per eye. The iCare IOP measurements were compared to those acquired on the same visit using Goldmann applanation tonometry to verify they did not differ significantly. The IOL Master (Zeiss Meditec, Dublin, CA) was used to measure the axial lengths and corneal curvatures of each eye. The curvature values were input to Spectralis to calibrate the magnification. The Spectralis was set to high resolution and auto-brightness mode. At each imaging session, three consecutive OCT image volumes, consisting of 24 radial scans (ONH-RC scan) equally spaced circumferentially around the ONH, were acquired using enhanced depth imaging mode for each eye. To optimize the image quality of the scans, the visible tissue was centered such that it occupied the lower two-thirds of the window and its brightness was adjusted until it reached the maximum. We refer to the anterior-posterior direction as the Z-direction, the radial direction as the R-direction, and the circumferential direction as the ϴ-direction. The in-plane resolution of each scan (RZ plane) was 768 × 495 pixels, which translated to 5.17 – 6.25 μm in R and 3.87 μm in Z. At Bruch’s membrane opening, the out-of-plane resolution was approximately 108 μm in Θ.

### Image segmentation

Each of the 24 radial scans was segmented by hand using FIJI.^15^ Within each OCT scan, the end of Bruch’s membrane opening (BMO) was marked on either side of the ONH and the anterior border of the LC was marked where visible between either side of the BMO (Figure 1). The anterior LC markings were fit to a continuous line using piecewise linear interpolation, and a parallel curvilinear border was drawn 250 μm posterior to the anterior border to represent the posterior LC, since the actual posterior limit of the LC is typically not visible in OCT images. The LC region was defined as the area between the two LC borders and within BMO delineated by vertical lines. LC thickness of 250 μm is based on our past microscopic measurements of the human LC.^17^

**Figure 1:**
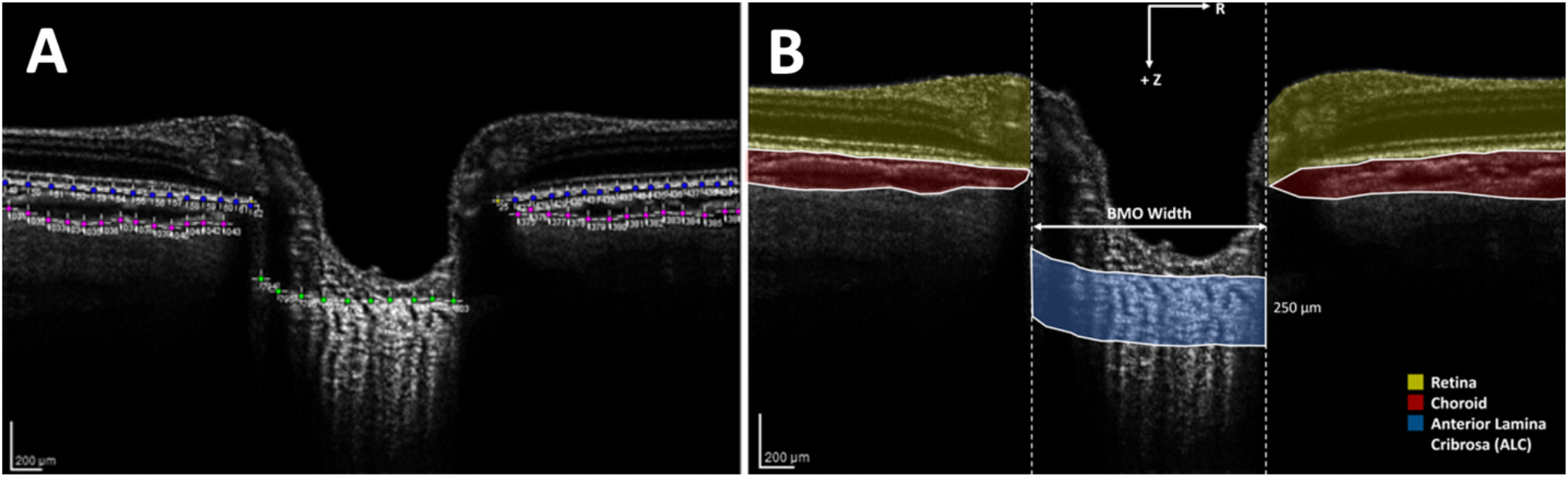
A) Segmentation of slice 1 of the 24 OCT images of the ONH. B) The segmented regions including the retina, choroid, BMO, and anterior LC. The anterior LC represents an area of 250 μm below the marked LC surface.

### Digital volume correlation and strain calculations

Prior to processing with digital volume correlation (DVC)^18^, the speckle pattern in OCT images was enhanced with contrast-limited adaptive histogram equalization (CLAHE) in FIJI. A gamma correction of 1.75 was then used to reduce noise in areas of low signal. Images were next imported into MATLAB 2019a (Mathworks, Natick, MA) to produce a 3-dimensional 3D matrix of 8-bit intensity values. DVC was applied to OCT image pairs before and after IOP change to estimate the 3D displacement field. These were smoothed to calculate the components of the Green-Lagrange strain tensor from the gradient of the displacement field as described in previous work.^15^ The three 3 normal strains components, *E*_*zz*_, *E*_*rr*_, and *E*_*θθ*_, indicate local expansions (positive) or contractions (negative) of the tissue in the thickness *(z)*, radial *(r)*, and circumferential *(θ)* directions. The three shear strain components *E*_*rz*_, *E*_*rθ*_, and *E*_*θz*_, denote local angular distortions in their respective planes. For example, *E*_*θz*_ for a cylinder results from a twist and *E*_*rθ*_ indicates transformation of the circular cross-section into an elliptical cross-section. From these strain components, we calculated the maximum principal strain (*E*_*max*_) and maximum shear strain (*Γ*_*max*_) in the RZ plane of the radial OCT images. We also divided strain data into the central and peripheral lamina, using half the radius of the LC outer border as separation. In addition, we divided the LC strain data by quadrant, using an X-shaped delineation to segregate superior, temporal, inferior and nasal quadrants.

In addition to LC strains, we measured the change in position of the anterior LC border from its displacement (*U*_*z*_) relative to the position of the BMO. LC border change was calculated as an average displacement across the entire LC border.

For each eye, we estimated the baseline displacement and strain errors by applying DVC to consecutive OCT image volumes acquired approximately 2 minutes apart of the same optic nerve head at the baseline IOP.^15^ Since the consecutive images were acquired under nominally the same IOP, non-zero values of strains and displacements denote error caused by the imaging conditions, image quality, and the imaging correlation algorithm. DVC fails or produces high baseline errors in regions of low contrast, such as where the LC is shadowed by overlying blood vessels. These regions excluded from subsequent strain analyses by filtering out points where the correlation coefficient was below 0.055.

The resulting average area of accurate DVC correlation was 51.1 ± 21.4% across the 23 image sets. The average baseline displacement error averaged over the LC were 0.016 ± 0.201μm for *U*_*z*_, 0.065 ± 0.267μm for *U*_*r*_, and 1.43 ± 4.25μm for *U*_*θ*._ The average baseline strain errors were 0.037 ± 0.118% for *E*_*zz*_, -0.020 ± 0.061% for *E*_*rr*_, -0.019 ± 0.150% for *E*_*θθ*_, 0.022 ± 0.102% for *E*_*rθ*_, 0.018 ± 0.077% for *E*_*rz*_, and - 0.029 ± 0.266% for *E*_*θz*_.

### Statistical analysis

We used paired and unpaired t-tests, as appropriate to determine the significance of the mean LC strains compared to no change and to baseline and correlation errors. Relationships between strains and other parameters including age, IOP, LC border movement, and glaucoma damage (MD, VFI, and RNFL thickness) were analyzed using analysis of variance and random effects multivariable regression models for the data from the reduced IOP group. For the unchanged IOP group, there was only one pair of eyes from one of the patients, so both eyes were included in the analysis of this group of 6 total eyes. For all statistical analyses, a p-value of 0.05 or less was considered significant.

## Results

### Intraocular pressure and lamina cribrosa depth changes

One week after medication change, the reduced IOP group had a mean IOP change of -5.8 ± 1.5 mmHg (p≤0.0001, paired t-test, 17 eyes of 12 patients), representing a mean decrease from baseline of - 29.6% ± 8.2%. The range of IOP change in this group was -3 to -8 mm Hg. Mean IOP change in the unchanged IOP group was -0.3 ± 0.8 mmHg (p=0.36, paired t-test, 6 eyes of 5 patients), or -2.7% ± 8.1%, which was insignificantly below baseline.

The anterior lamina depth (ALD) change from baseline after IOP change was defined as positive if the anterior LC surface moved posteriorly (away from the cornea) and negative if it moved anteriorly (towards the cornea). For reduced IOP eyes, mean ALD change was 2.05 μm (95% confidence interval (CI)= (−0.02, 4.1), random effects model accounting effects of inclusion of both eyes for some patients; p=0.052 compared to zero change). Thus, there was only a small mean posterior movement of the LC, with some eyes moving posteriorly and some anteriorly. ALD change was not significantly related to IOP change nor to percent IOP change (p=0.94, random effects model). In the unchanged IOP group, mean ALD change was +1.50 ± 2.27 μm, but not significantly different from zero change (p=0.17, paired t test).

### LC strains

For the reduced IOP group, the LC strain response to IOP decrease were significantly larger than the baseline (reproducibility) error for *E*_*zz*_ and *E*_*rr*_ strains (p=0.002, 0.011, respectively, random effects models) (Table 1, Figure 2). The normal strain in the thickness direction *E*_*zz*_ = 1.0% ± 1.1% was tensile (greater than zero, p = 0.003) and the radial strain *E*_*rr*_ was compressive, -0.3% ± 0.5%; p=0.012). In addition, the maximum principal (tensile) strain and maximum shear strain in the reduced IOP group were significantly greater than zero: *E*_*max*_ = 1.7% ± 1.0% and *Γ*_*max*_ = 1.4% ± 0.7%; (both p<0.0001).

**Table 1:** Lamina cribrosa strains in the reduced IOP and unchanged IOP groups. Mean strains ± standard deviation (strains are unitless). N=17 eyes in reduced IOP group and 6 eyes in unchanged IOP group.

|  | Reduced IOP | P-Value | Unchanged IOP | P-Value |
| --- | --- | --- | --- | --- |
| $E_{zz}$ | $0.010 \pm 0.011$ | **0.003** | $0.003 \pm 0.003$ | 0.054 |
| $E_{rr}$ | $-0.003 \pm 0.005$ | **0.012** | $0.001 \pm 0.009$ | 0.76 |
| $E_{\theta\theta}$ | $-0.001 \pm 0.009$ | 0.70 | $-0.002 \pm 0.009$ | 0.76 |
| $E_{r\theta}$ | $0.002 \pm 0.008$ | 0.35 | $-0.011 \pm 0.017$ | 0.20 |
| $E_{\theta z}$ | $-0.001 \pm 0.005$ | 0.32 | $0.001 \pm 0.006$ | 0.65 |
| $E_{rz}$ | $0.000 \pm 0.006$ | 0.77 | $-0.001 \pm 0.004$ | 0.63 |
| $E_{max}$ | $0.017 \pm 0.010$ | **<0.0001** | $0.013 \pm 0.012$ | 0.053 |
| $\Gamma_{max}$ | $0.014 \pm 0.007$ | **<0.0001** | $0.011 \pm 0.009$ | **0.041** |

**Figure 2:**
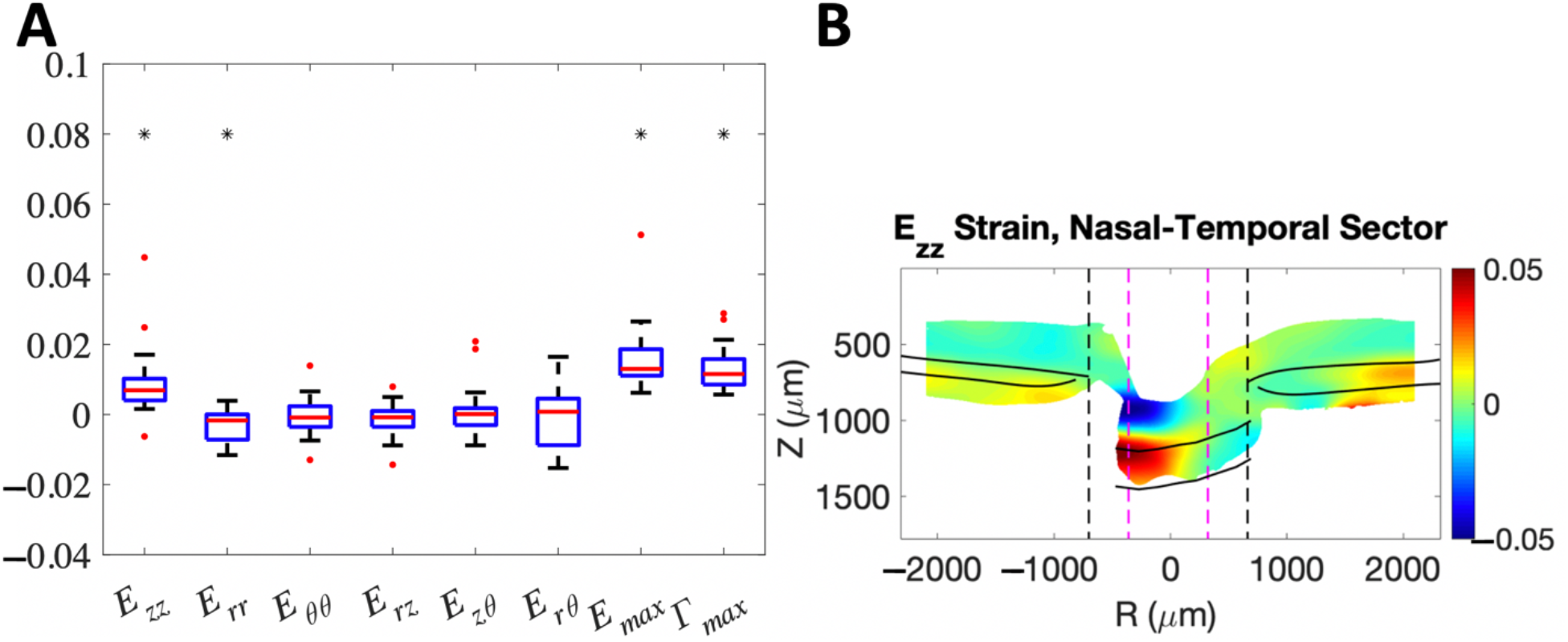
A) Strains in reduced IOP group (n=17 eyes) where asterisk (*) indicates p-value ≥ 0.05. B) Example of strain map for *E*_*zz*_ strain for a radial scan along the inferior-superior direction for representative eye from reduced IOP group (IOP 7 mm Hg lower than baseline), showing a regionally varying tensile strain (red region, thickening) in the LC.

For the unchanged IOP group, which served here as a form of control group, strains had only borderline significant differences from zero in *E*_*zz*_, *E*_*max*_, and *Γ*_*max*_ (p=0.054, 0.053, 0.041, respectively). The mean *E*_*zz*_ for this group was 0.3% ± 0.3%, 3 times smaller than that of the reduced IOP eyes and not significantly greater than baseline error (p=0.08).

There were no significant differences in either group in *E*_*zz*_ or *E*_*max*_ strains in the central compared to the peripheral LC (all p>0.63 for reduced IOP group; all p>0.34 for unchanged IOP group), nor did strains differ significantly by LC quadrant (all p>0.65 for reduced IOP group; all p>0.66 for unchanged IOP group).

### LC strains in reduced IOP group compared to other variables

In the reduced IOP group, strains *E*_*max*_ and *Γ*_*max*_ were significantly larger with greater % IOP decrease while *E*_*rr*_ was significantly more negative with greater % IOP decrease (for % decrease: <0.0001, <0.0001, and p=0.048, respectively, random effects models; Figure 3). *E*_*max*_ and *Γ*_*max*_ strains were also significantly associated with the magnitude of IOP decrease (p=0.0029, 0.0020, respectively, random effects models). All strains were unrelated to patient age (all p≥0.088). Among all strains, only *E*_*rr*_ was significantly related to ALD change, becoming more negative with greater posterior ALD change (p=0.015). Strain data were compared to the degree of glaucoma damage in visual field and RNFL thickness. Strains were not significantly related to either MD or VFI indexes in field data (all p≥0.14). Strains *E*_*zz*_, *E*_*max*_, and *Γ*_*max*_ were larger as RNFL thickness increased in models (all p<0.001). This outcome appeared to be due to 2 outlying values with higher RNFL thickness (Supplemental data).

**Figure 3:**
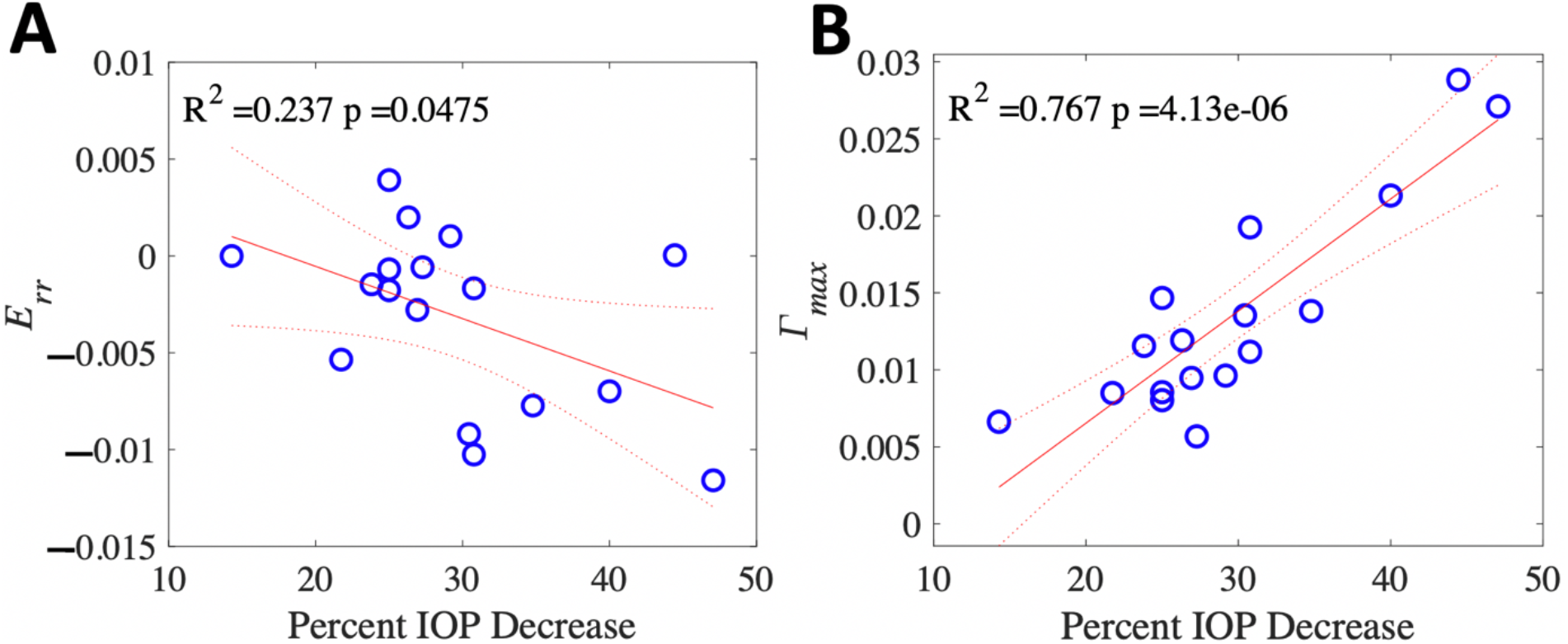
A higher percent decrease in IOP was associated with A) more radial contraction (negative *E*_*rr*_) and B) larger maximum shear strain (*Γ*_*max*_). Data from reduced IOP group (n=17 eyes). Linear regression analysis.

Since we determined that strains were related to the degree of IOP lowering, we calculated for each strain response the value called compliance, defined as the strain per mmHg IOP decrease. In random effects models, compliances of strains *E*_*zz*_, *E*_*max*_, and *Γ*_*max*_ were significantly larger with more abnormal MD index value in the visual field (p=0.02, 0.001, 0.011, respectively, Figure 4). Likewise, compliances for *E*_*max*_ and *Γ*_*max*_ were larger with worsening VFI (p=0.015, 0.010, respectively). Compliances for the same 3 strains (*E*_*zz*_, *E*_*max*_, and *Γ*_*max*_) were significantly related to RNFL thickness, but when the 2 outlying values were omitted from the analysis there was no demonstrable relation of strains to RNFL thickness.

**Figure 4:**
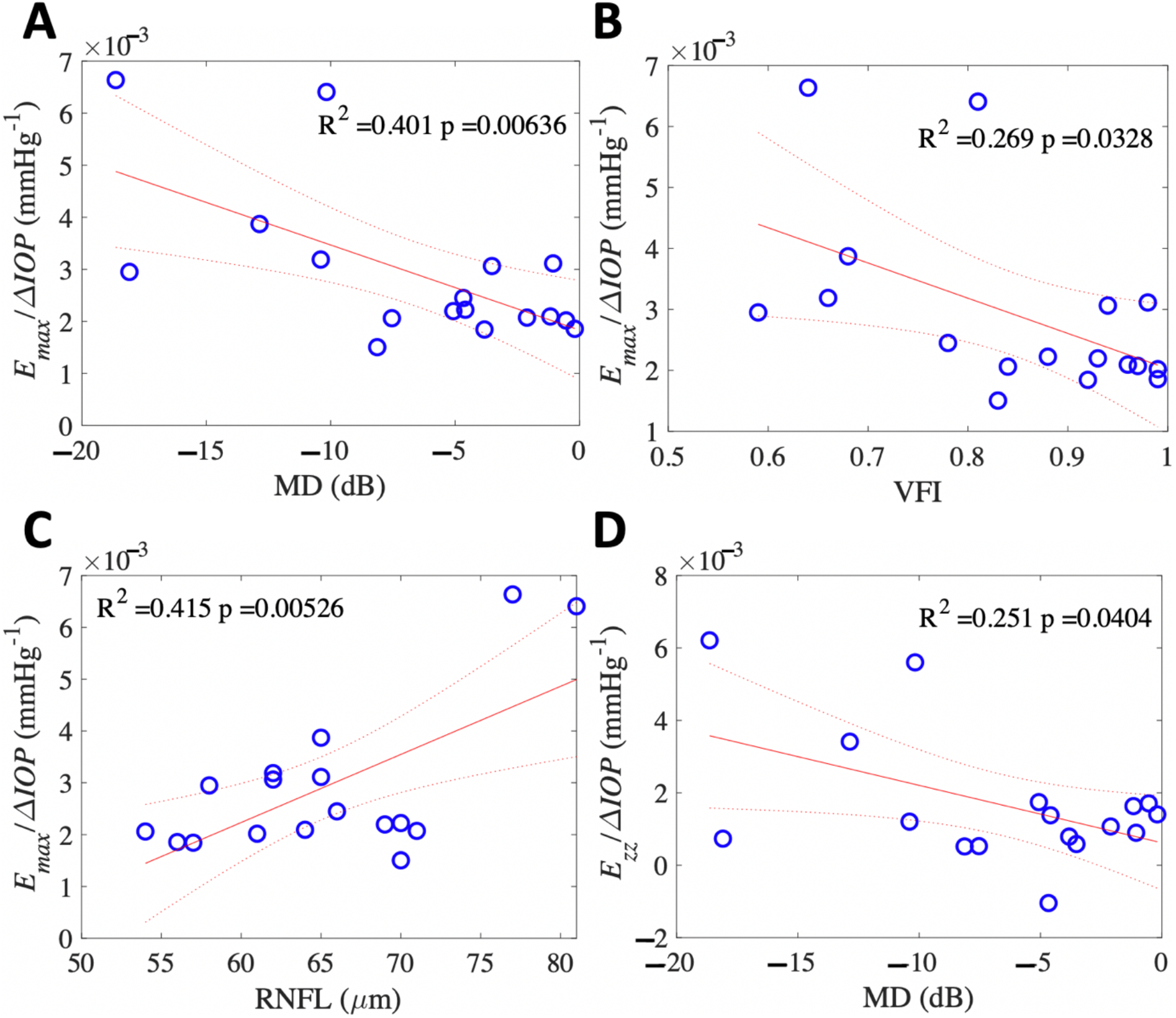
Eyes with greater glaucoma progression as indicated by A) more negative MD and B) lower VFI had greater compliance of maximum principal strain (*E*_*max*_/ΔIOP) indicating a softer strain response. C) However, a thicker RNFL thickness was associated with greater compliance of *E*_*max*_. D) A more negative MD was associated with greater compliance of *E*_*zz*_. Data from reduced IOP group (n=17 eyes). Linear regression analysis.

## Discussion

These data confirm our previous findings that reduction of IOP leads to measurable strains in the LC with sequential OCT imaging. There are several similarities to our initial reports. In Czerpak et al.^15^, we imaged eyes before and 20 minutes after post-operative laser suturelysis in eyes that had undergone trabeculectomy. The patients in that report had somewhat greater structural and functional damage (mean MD = -11.5 ± 8.9 dB) than the eyes in this dataset (for reduced IOP group: mean MD = - 6.6 ± 5.8 dB), and the suturelysis eyes had greater mean IOP lowering. Despite these differences, the strains measured one week after changes in eye medication were of similar magnitude to those immediately after suturelysis. This suggests that after IOP lowering the LC exhibits viscoelastic behavior with strain increasing more at one week than after 20 minutes. One group found that experimental IOP increase by 2 minutes of ophthalmodynamometry pressure on the eye has also demonstrated measurable strains, but with dramatically greater IOP elevations.^9^ Our findings show that a simple change in eye drop medication produces strains in the LC that are assessable at a typical follow-up visit. There may be an advantage to methods of IOP lowering for which strain can be measured after at least one week in order to produce more easily measurable strain values.

Furthermore, these data provide further evidence that eyes with greater visual field damage have greater, not less, LC strain. We^15^ and others^16^ have previously found that eyes exhibiting worse injury have a more compliant LC. In fact, we also found similarly in study of control and glaucoma eyes *post mortem* that eyes with worse histological axon loss had greater compliance than mildly damaged eyes.^19,20,21^ These findings seem to contradict prior conclusions that glaucoma eyes are stiffer after suffering damage. Prior research, however, did not routinely measure LC strains, but rather indirect measures of portions of the sclera more distant from the peripapillary area.^22,23^ We previously measured the LC strain in *ex vivo* inflation tests and found that the LC strain response of mildly damaged glaucoma eyes was stiffer than those of undamaged normal eyes.^21^ The most relevant zones for glaucoma damage are in the immediate ONH region. Conceivably, the greater strains we have estimated in the LC with worse damage could result from greater glaucoma damage causing remodeling in which LC beams become thinner.^24^ Indeed, LC curvature in inflation testing was greater with greater damage, implying a more compliant response.^25^ Both monkeys with chronic IOP elevation and human glaucoma eyes are reported to have thinner than normal LC, suggesting more compliant not stiffer behavior.^26,27^ The peripapillary sclera in experimental and human glaucoma has a less ordered circumferential fiber pattern^28,29^ that may behave more rigidly, intensifying the effect of the translaminar pressure gradient to cause greater LC strain. Indeed, we have estimated posterior scleral strains using the OCT method described here and find that scleral strains are proportionately smaller than the LC of the same eye, and that the strains in the two zones are significantly related to each other.^30^ We must keep in mind that our data are cross-sectional and do not indicate what the strain state of damaged glaucoma eyes was at baseline.

The medication change approach produced not only similar magnitudes of strains, but strain change in the same directions as our previous data after suturelysis. IOP decrease in both paradigms caused the LC to expand in thickness (*E*_*zz*_ strain) and to contract in radius (*E*_*rr*_ strain). One could visualize this change as if the LC were a container that changed shape from a tuna fish can to an orange juice can. In both studies, larger IOP decreases led to more tensile *E*_*zz*_ and greater maximum principal (*E*_*max*_) and maximum shear strains (*Γ*_*max*_). Likewise, there were consistent findings that greater compliance of *E*_*max*_ and *Γ*_*max*_ were associated with worse MD and VFI visual field indexes. In both situations, the ALD change was minimal in this time frame.

Interestingly, the single difference with our previous study was the fact that greater strain was associated with thicker average RNFL in this study, but thinner RNFL in our suturelysis data. However, this result deserves further study with greater numbers of eyes, as it appears to have been generated largely by 2 outlying values. It is well-known that RNFL thickness more rapidly declines than visual field sensitivity in glaucoma. In our previous study, the vast majority of eyes had RNFL average thickness values lower than 65 μm, while in the present study, the majority had RNFL thickness greater than 65 μm. By contrast, the visual field indices are a more continuous reflection of glaucoma damage, while the RNFL data decline earlier and have a floor effect at 50 μm. The seeming difference in our two studies related to structural glaucoma damage related to strains may actually, therefore, be another indication that stage of glaucoma damage is related to LC strain behavior.

The validity of these OCT strain measurements was further confirmed by the finding that strains exceeded the reproducibility error. Furthermore, in dividing our test subjects into those whose IOP did not change with medication alteration and those that did, we could show that strains were largely insignificant when IOP was not different from baseline.

While our data had significant associations with the present sample, it will be important to substantially expand the dataset to confirm several important hypotheses. Further studies should include imaging of non-glaucoma control persons who would be consented to take eyedrops to lower IOP. These should include both fully normal, age-matched persons, as well as ocular hypertensive eyes with no damage. The expansion of the sample size could also allow estimation of the potential effects on strain of other variables, such as axial length, refraction, sex, and race. Our statistical method accounted for the inclusion of both eyes of some patients, but further work should expand the bilateral testing to explore the possibility that an eye that is more damaged, or more likely to be damaged in the future, has different strain behavior.

In conclusion, the use of DVC analysis of OCT images before and after IOP change is a feasible means for measuring *in vivo* LC strains. The strain measures were confirmed as greater with worse visual field damage, suggesting that with further research such imaging may provide a biomarker for glaucoma susceptibility at routine clinic visits. Longitudinal studies are merited to investigate the biomechanical response to IOP change over time in these patients.

## Data Availability

All data produced in the present study are available upon reasonable request to the authors

## Abbreviations

LC: lamina cribrosa
ONH: optic nerve head
OCT: spectral domain optical coherence tomography
RNFL: retinal nerve fiber layer
MD: mean deviation
VFI: visual field index
IOP: intraocular pressure
dB: decibels

