## Supplemental Figure S1 for "A noninvasive clinical method to measure *in vivo* mechanical strains of the lamina cribrosa by optical coherence tomography"

**Supplemental Material**

In relating the strains in the reduced IOP group to the average RNFL values, there were two eyes that differed dramatically from the other 15 eyes, in that the RNFL values were 14% higher than any of the others (despite their visual field parameters indicating moderate damage in one eye and severe damage in the other eye) and whose compliance values were also more than twice as high as any of the other eye data (Figure). Since the RNFL values of these two eyes were not compatible with their visual field findings, we conducted a second analysis without these two outliers. This showed that there was no significant relationship between strain compliance and average RNFL thickness in the remaining 15 eyes. The linear regression relationship was insignificant (p=0.96; r2 = 0.0002).

Figure S1: Removing two RNFL values (top right) that were not compatible with their visual fields, RNFL thickness was not associated with compliance of $E_{max}$.

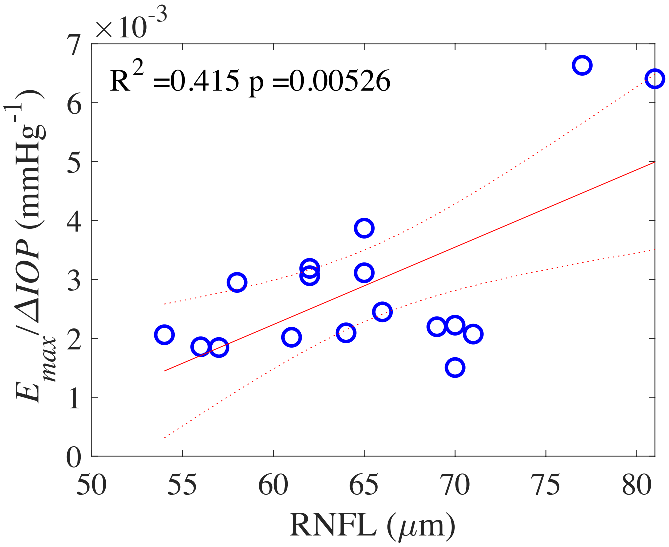
